# GIS-Based Analysis Framework for identifying COVID-19 Incidence and Fatality Determinants at National Level Case study: Africa

**DOI:** 10.1101/2021.01.12.21249661

**Authors:** Mahmoud A. Hassan, Ramy Mohamed Ghazy, Rofida G. Abdelwahab, Toka A. Elbarky

## Abstract

**Background:** COVID-19 pandemic is an extraordinary threat with significant implications in all aspects of human life, therefore, it represents the most immediate challenges for all countries all over the world.

**Objectives:** This study is intended to develop a GIS-based analysis model to explore, quantify and model the relationships between COVID-19 morbidity and mortality and their potential predictor variables.

**Method:** For this purpose, a model was developed to estimate COVID-19 incidence and fatality rates in Africa up to 16^th^ of August 2020 at the national level. The model involved Ordinary Least Squares (OLS) and Geographically Weighted Regression (GWR) analysis through ArcGIS was applied.

**Result:** Spatial Autocorrelation Analysis revealed that there was positive spatial autocorrelation in COVID-19 incidence (Moran index 0.16. *P* value <0.1), and fatality (Moran index 0.0.35, *P* value<0.01) rates within different African countries. At continental level, OLS revealed that COVID-19 incidence rate was found to be positively associated with overcrowding, health expenditure, HIV infections and air pollution and negatively associated with BCG vaccine (β=2.97,1.45, 0.01, 3.29, −47.65 respectively, *P*< 0.05) At the same time, COVID-19 fatality was found to be positively related to asthma prevalence and tobacco use. Yet, certain level of inconsistency was noted in the case of COVID-19 fatality, which was negatively related to elder population, poverty, and cardiovascular mortality (*P*<0.05). This model showed convenient level of validity in modeling the relationship between COVID-19 incidence as well as fatality and their key predictors using GWR. In this respect, the model explained about 58% and 55% of the variance in COVID-19 incidence and fatality rates, respectively, as a function of considered predictors.

**Conclusion:** Application of the suggested model can assist in guiding intervention strategies, particularly in case of local and community level whenever the data on COVID-19 cases and predictors variables are available.

## Introduction

COVID-19 is the key global challenge nowadays with a lockdown of more than6 billion people (Iqbal et al., 2020). The novel coronavirus (SRAS-COV-2) was firstly, discovered at the end of 2019 in Wuhan, China after causing a cluster of pneumonic cases. In February 2020, the World Health Organization (WHO) designated the disease as COVID-19, which stands for coronavirus disease 2019 (WHO, 2020c). In fact, this virus spread rapidly resulting in an epidemic throughout China, followed by a massive spread across the world. On March 11, 2020, WHO declared a COVID-19 as a pandemic disease due to the increasing number of cases in large number of countries all over the world at rapid rates and the severity of disease (García-Basteiro et al., 2020). Up till 22^nd^ of November 2020, globally, more than 57 million individual caught infection with more than 1.3 million confirmed deaths. In Africa, more than 2 million of COVID-19 cases and about 49 thousands confirmed deaths were reported up to date (WHO, 2020b).

The transmission of COVID-19 is primarily transmitted through direct person-to-person transmission. Furthermore, it has been detected in non-respiratory specimens (stool, blood, ocular secretions, and semen), yet the role of these sites in transmission is uncertain (ATCTW, 2020). More recently, the airborne transmission was reported as a probable transport pathway to spread COVID-19 (Pani et al., 2020). It is worthy to mention that, asymptomatic individual can transmit infection (Rothe et al., 2020). Symptomatic patient infectiousness started 2.3 days before symptoms onset (peak 0.7 days before symptom onset) and declined within seven days. After a week to 10 days, the risk of diseases transmission is unlikely, chiefly for immunocompetent patients with non-severe infection. Broadly speaking, degree of closeness and duration of contact increases the risk of infection (He et al., 2020).

For combating and proper management of COVID-19 spread, there is a need to understand the key determinants of its incidence. Therefore, a considerable number of research work was undertaken to examine various determinants of COVID-19 incidence and fatality. Generally, <u>weather conditions</u> were suggested to have impacts on COVID-19 incidence, where wet and cold conditions assumed to contribute to the rapid spread of COVID-19. In this respect, it was found that COVID-19 cases and deaths are negatively correlated with temperature (Iqbal et al., 2020) and positively correlated with absolute humidity (Pani et al., 2020). Also, it was argued that that air pollution can promote to sustained transmission of COVID-19, where COVID-19 incidence was found to be positively correlated with PM2.5 and NO2 (Li et al., 2020). This may highlight the role of <u>quality of life</u> (QOL) as one of the main determinants of COVID-19 incidence. In addition to air pollution, QOL can be manifested in access to drinking water and sanitation services.

Also, the association between Particulate Matter (PM) pollution and COVID-19 case fatality rate was emphasized (Yao et al., 2020). Moreover, it was argued that existing <u>health security capacities</u> are essential for tackling public health risks including infectious disease outbreaks such as COVID-19. in this respect, it was found that half of world countries have not sufficient operational readiness capacities for dealing with potential health emergencies associated with COVID-19 (Pung et al., 2020).

Furthermore, vulnerability of individuals to COVID-19 morbidity and mortality can be increased by <u>comorbidity of some chronic diseases</u>. For example, it was found that elderly individuals (65 years old and above), who have comorbidities, such as hypertension or diabetes mellitus (DM), are expected to develop a more severe course and progression of the COVID-19 (Sanyaolu et al., 2020). Such an argument may imply also the role of age structure in COVID-19 incidence, where elder population are likely to catch COVID-19. As for <u>socioeconomic and demographic conditions</u>, a wide range of determinants were examined such as age structure (Baron, 2020; Goldstein and Lee, 2020; Omori et al., 2020), which was found to have a significant impact of COVID-19 fatality. Accordingly, the proportion of elder population (65 years or above) was suggested to be a predictive for COVID-19 fatalities (Hawkins et al., 2020). Meanwhile, the case-fatality ratio (CFR) was found to be correlated significantly with population density (Baron, 2020). Also, it was argued that the COVID-19 fatality rates were found to be associated with poverty and urban population proportions (Finch and Hernández Finch, 2020). Both COVID-19 cases and fatalities were found to strongly associated with lower education levels (Hawkins et al., 2020) and tobacco smoking (Baron, 2020).

Many countries have adopted the national lockdown, limiting travel, and spatial rather than “social” distancing to control the local transmission of COVID-19 (Bi et al., 2020; Tian et al., 2020). Such strategies were found to be effective to mitigate COVID-19 transmission (Kraemer et al., 2020; Zhao et al., 2020). Among these strategies for mitigating transmission, limiting travel was found to have modest effect while transmission reduction measures like lockdown showed higher levels of effectiveness in the disease transmission control (Chinazzi et al., 2020). This generally implies that <u>population mobility</u> is one of the main determinants of COVID-19 incidence.

Mapping disease outbreaks can assist in understanding their key determinants. In this respect, thematic maps may be misleading in predicting the spatial pattern of the diseases. Alternatively, analyzing spatial pattern is more effective as it implies assessing statistical significance of spatial clustering or dispersion, mapping clusters, if any, and modeling spatial relationships (Fischer and Getis, 2010). Accordingly, the key factors contributing to the diseases incidence and fatality can be determined.

Such an overall objective can be attained through the following three specific objectives including: analyzing spatial variations of COVID19 cases and delineating the main hotspots, exploring various determinants of COVID-19 spatial pattern. and identifying the most significant determinants of COVID-19. Such a GIS-based model can provide better understanding of the key factors underlying COVID-19 incidence and fatality, which, in turn, may guide intervention measures in the future.

## Methodology

To identify the key determinants of COVID-19 incidence and fatality, a methodology of four main steps was developed (Figure 1) as follows:

**Figure 1:**
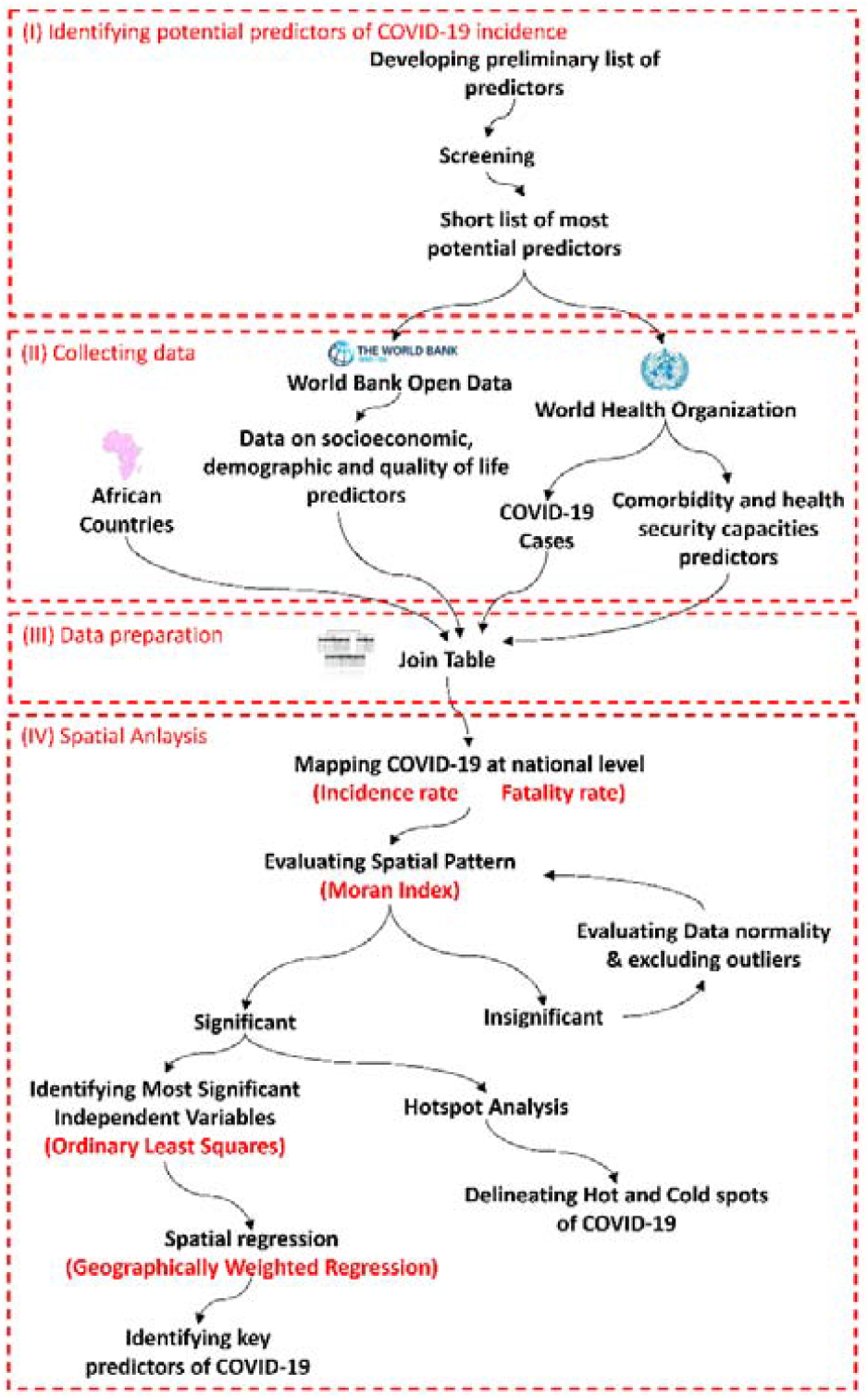
Analysis framework for identifying COVID-19 incidence and fatality determinants through GIS

- Identifying potential predictors of COVID-19 incidence: Firstly, a preliminary list of COVID-19 incidence predictors was developed. The list of identified predictors reflected 33 of COVID-19 incidence predictors. Thereafter, these preliminary predictors were screened in accordance to some criteria such as relevance and data availability, therefore, a short list of 14 predictor variables of COVID-19 incidence was developed that reflect different categories of predictors including: demographic and socioeconomic conditions in addition to comorbidity, health security capacities, quality of life and population mobility (Table 1). Meanwhile, a short list of 16 predictor variables of COVID-19 fatality was developed that reflect different categories of predictors including: demographic and socioeconomic conditions in addition to comorbidity and health security capacities (Table 2Table 1).
- Collecting data on COVID-19 and potential predictors: Data on total confirmed cases and deaths at national level in Africa was up to 16^th^ of August 2020 were acquired from World Health Organization (WHO) portal, which provides data on the global, regional and country-level COVID-19 cases and deaths in regular basis (WHO, 2020b). Meanwhile, data on most potential predictor variables were acquired form The World Bank Group (The World Bank Group, 2020) and The Global Health Observatory (WHO, 2020a).
- Data preparation: This involved developing a geodatabase for African countries including a key polygon feature class representing the territories of African countries with their main attributes. Moreover, collected data on various predictors were tabulated and COVID-19 incidence and fatality rates were calculated. Thereafter, data on COVID-19 incidence as well as potential predictors were integrated into the developed geodatabase through join table.
- Spatial analysis: This involves mapping COVID-19 incidence and evaluating the spatial pattern of COVID-19 incidence through Spatial Autocorrelation Tool (Moran’s I index). Based upon the results of Moran index, the spatial pattern of COVID-19 incidence was evaluated and outlier records, whether statistically or spatially, are excluded. Thereafter, the main hot spots were delineated through Hot Spot Analysis. Meanwhile, the relationships between potential predictors as independent variables and COVID-19 incidence or fatality as dependent variable were calibrated and most significant independent variables are identified through Ordinary Least Squares Tool. Finally, Geographically Weighted Regression (GWR) is applied to explore the predictive power of such variables locally for each county, as it is not expected that Covid-19 driving factors would be uniform across different countries with varied conditions. GWR was initially introduced by (Brunsdon et al., 1996) to extend the traditional regression analysis for modeling local relationships between a certain phenomenon that is varied over space and a number of predictors so that the coefficients in the model rather than being constant are location-specified. Due to its analytical power, GWR was recently applied for modeling and explaining the spatial relationships in a variety of fields (Cahill and Mulligan, 2007; Huang and Leung, 2002; Mennis, 2013)

**Table 1:** List of potential predictors of COVID-19 incidence

| Categories | Predictors | Indicators |
| --- | --- | --- |
| Socioeconomic and demographic predictors | Overcrowding | Population Density (person/Km<sup>2</sup>) |
|  | Educational level | Illiteracy rate (%) |
|  | Income | GDP per capita |
|  | Poverty incidence | Proportion of the population living in extreme poverty (%) |
| Comorbidity predictors | Vaccine protects against tuberculosis | Proportion of one-year old population immunized (%) |
|  | HIV infections | Rate of HIV infections (per 1000 uninfected population) |
|  | Tuberculosis incidence | Rate of Tuberculosis incidence (per 100 000 population) |
|  | Tobacco use | Prevalence of tobacco use among persons 15 years and older (%) |
| Health security capacities predictors | Health expenditure | Domestic general government health expenditure (GGHE-D) as percentage of general government expenditure (GGE) (%) |
|  | Stringency index | Government Response Stringency Index |
| Quality of life | Using a hand-washing facility | Proportion of population using a hand-washing facility with soap and water (%) |
|  | Access to drinking water services | Proportion of population using at least basic drinking water services (%) |
|  | Access to sanitation services | Proportion of population using at least basic sanitation services (%) |
|  | Air pollution | Annual mean concentrations of fine particulate matter (PM2.5) in urban areas (µg/m<sup>3</sup>) |

**Table 2:** List of potential predictors of COVID-19 fatality

| Categories | Predictors | Indicators |
| --- | --- | --- |
| Socioeconomic and demographic predictors | Age Structure | Median age (year) |
|  | Elder population | Proportion of elderly Population (Above 65 years old) (%) |
|  | Human Development | Human development index |
|  | Poverty | Proportion of the population living in extreme poverty (%) |
| Comorbidity predictors | Diabetes mellitus prevalence | Proportion of diabetic population (ages 20 to 79) (%) |
|  | Cardiovascular fatality | Death rate from cardiovascular disease (per 100,000 people) |
|  | Asthma prevalence | Proportion of asthmatic population (%) |
|  | Cancer prevalence | Proportion of population Cancer (%) |
|  | Human immune deficient virus (HIV) infections | New HIV infections (per 1000 uninfected population) |
|  | Tuberculosis incidence | Tuberculosis incidence (per 100 000 population) |
|  | Tobacco use | Prevalence of tobacco use among persons 15 years and older (%) |
| Health security capacities predictors | Density of physicians | Density of medical doctors (per 10 000 population) |
|  | Density of nursery | Density of nursing and midwifery personnel (per 10 000 population) |
|  | Density of pharmacist | Density of pharmacists (per 10 000 population) |
|  | Health expenditure | Domestic general government health expenditure (GGHE-D) as percentage of general government expenditure (GGE) (%) |
|  | Hospital beds ratio | Hospital beds per 1,000 people |

## Results and Discussion

Up till 16^th^ of August 2020, African countries recorded 1.1 million confirmed cases of COVID-19 and more than 25,000 deaths. COVID-19 cases varied widely among different countries ranging between 583653 in the cases of South Africa and 285 in Eretria. Similarly, COVID-19 deaths ranged between 0 in Eretria and 11,677 in south Africa. As for COVID-19 incidence rate, it was found to be about 0.97 (per 100,000 population) on average ranging between 0.009 and 9.97 (Table 3). Such noticeable high variations, which indicates to uneven distribution of COVID-19 cases among different countries, can be actually attributed to the accuracy of registration procedures in these countries.

**Table 3:** Descriptive statistics of COVID-19 incidence and fatality rate in African countries

| Variables | Minimum | Maximum | Mean± Standard Deviation |
| --- | --- | --- | --- |
| COVID-19 Incidence rate<br>(per 100,000 of population) | 0.01 | 9.97 | 0.96±1.69 |
| COVID-19 fatality rate<br>(per 100,000 of population) | 0.00 | 0.20 | 0.02±0.03 |

To evaluate the distribution pattern of COVID-19 incidence and deaths rates, Spatial Autocorrelation analysis (Moran’s I index) was applied. Preliminary trials of Spatial Autocorrelation Analysis revealed that insignificant spatial autocorrelation in COVID-19 incidence and deaths rates due to existence of outlier records, whether spatially such as Madagascar, or statistically such as South Africa, Gabon and Djibouti. Therefore, these outliers were excluded. The results of Moran’s I index revealed that there was positive spatial autocorrelation in COVID-19 incidence and deaths rates within different African countries. It was found that the Moran’s I index for COVID-19 incidence rate recorded a small value (0.16), a relatively high z-score (1.67) and *P* value (<0.10) (Figure 2-a). Meanwhile, the Moran’s I index for COVID-19 deaths rate recorded a relatively higher value (0.35), z-score (3.16) and smaller *P* value (<0.01) (Figure 2-b), which indicate that the relationship is more significant in the case of deaths compared to confirmed cases.

**Figure 2:**
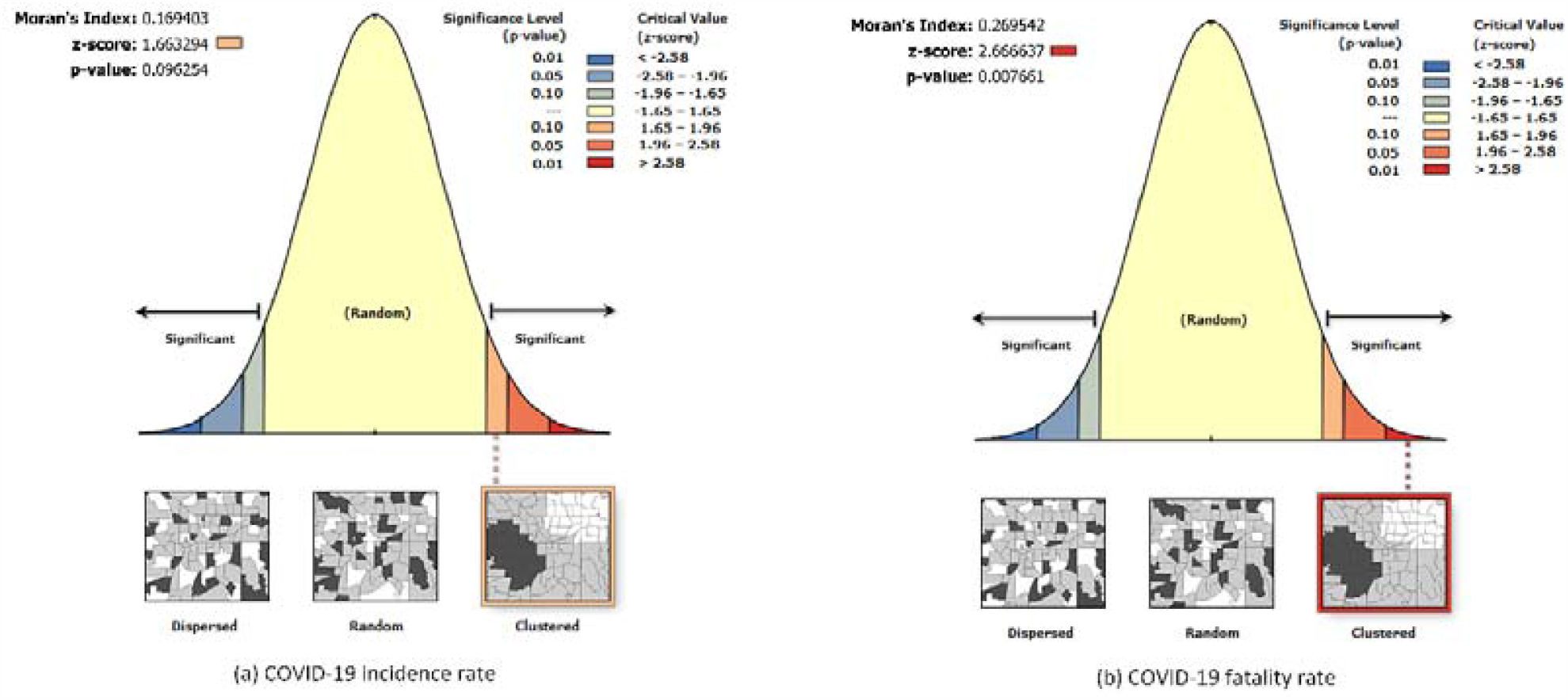
Spatial auto correlation analysis

Generally, it can be argued that, based on statistically significant *P*-value and high positive z-score, the spatial distribution of COVID-19 incidence and deaths rates tend to be clustered and the null hypothesis of Complete Spatial Randomness can be rejected. Hot Spot analysis revealed that a cold spot of COVID-19 incidence and fatality rates was noticeably identified in East of Africa (Figure 3-a & b). Meanwhile, three hot spots of COVID-19 incidence rate were identified. These hot spots included Egypt and Libya in the north east, Morocco, Mauritania, Senegal and Gambia in the north west in addition to Namibia in the south (Figure 3-a). Also, two hot spots of COVID-19 fatality rates were identified in the north-eastern and north-western parts of the continent (Figure 3-b) corresponding to but more significant compared to hot spots of incidence rate.

**Figure 3:**
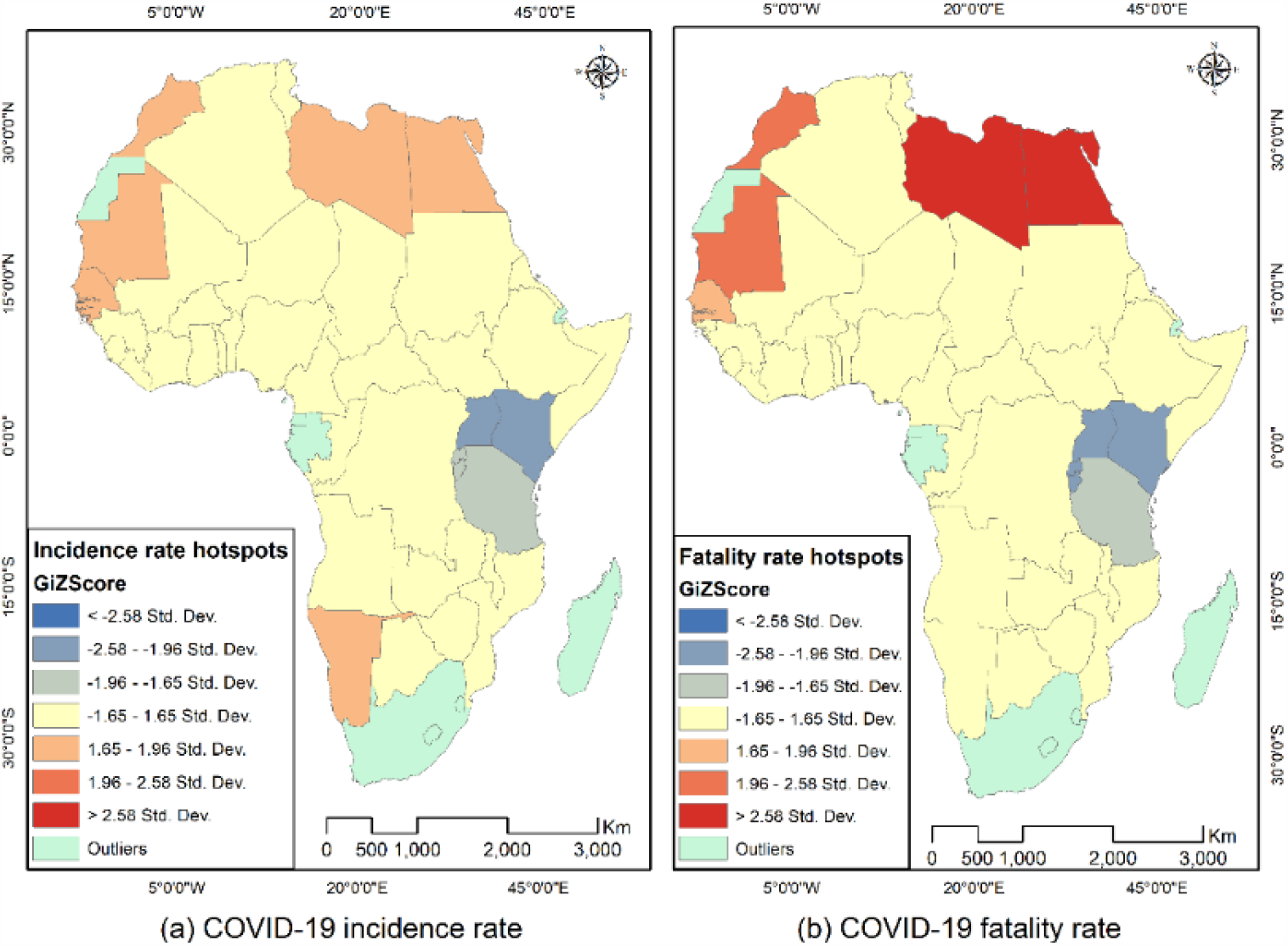
Hot spot analysis of COVID-19 incidence and fatality rates

To identify the key predictors of COVID-19 incidence and fatality rates, Geographically Weighted Regression (GWR) analysis through ArcGIS was applied. As a prerequisite for GWR, there would a need to evaluate multicollinearity among considered predictors (explanatory variables). For this purpose, a model was developed to estimate COVID-19 incidence and fatality rates in Africa up to 16^th^ of August 2020, at the national level, using v analysis. The relationships between the explanatory variables and COVID-19 incidence and fatality rates are assumed to be the same at various countries. As a result of OLS model, the relationships between COVID-19 incidence and fatality rates on one hand and various considered explanatory variables, on the other hand, were found statistically insignificant. Also. it was found that some variables have large value of Variance Inflation Factor (VIF), which indicate redundancy among these variables. Accordingly, through an iterative process, variables with large VIF value have been excluded step-wise and, the remaining explanatory predictor variables in terms of COVID-19 incidence included: overcrowding, vaccine against tuberculosis, HIV infection, health expenditure and air pollution. Meanwhile, the remaining explanatory predictor variables in terms of COVID-19 fatality included: elder population, poverty, cardiovascular fatality, asthma prevalence and tobacco use (Table 4).

**Table 4:** Ordinary Least Squares Regression Model

| Variable | Coefficient (β) | t-Statistic | Probability | VIF |
| --- | --- | --- | --- | --- |
| COVID-19 incidence |  |  |  |  |
| Intercept | 0.44514 | 1.97964 | 0.05543 | ----- |
| Overcrowding | 2.97288 | 4.31716 | 0.00012 | 1.08596 |
| Health expenditure | 1.45380 | 2.28912 | 0.02805 | 1.30737 |
| BCG vaccine | -47.65202 | -2.62772 | 0.01256 | 1.15719 |
| HIV infections | 0.01144 | 2.54531 | 0.01535 | 1.15583 |
| Air pollution | 3.28993 | 2.89638 | 0.00638 | 1.02464 |
| R<sup>2</sup> | 0.45351 |  |  |  |
| COVID-19 fatality |  |  |  |  |
| Intercept | 0.01512 | 2.07490 | 0.04520 | ----- |
| Elder population | -0.00258 | -2.23400 | 0.03178 | 2.07770 |
| Poverty | -0.00028 | -3.77480 | 0.00058 | 1.97820 |
| Cardiovascular fatality | -0.00031 | -2.66490 | 0.01146 | 1.90340 |
| Asthma prevalence | 0.00420 | 3.10450 | 0.00370 | 2.72620 |
| Tobacco use | 0.00005 | 2.94160 | 0.00568 | 1.16880 |
| R<sup>2</sup> | 0.53377 |  |  |  |

The results of OLS model showed that all explanatory variables of both COVID-19 incidence and fatality are statistically significant (*P* < 0.05). COVID-19 incidence rate was found to be positively associated with overcrowding (β=2.97), health expenditure β=1.45), HIV infection (β=0.01) and air pollution (β=3.29) and negatively associated with BCG vaccine (β=-47.65). These findings are consistent with expectations. The only exception was in the case of health expenditure that is supposed to be negatively related to COVID-19 incidence rate. This can be explained by the fact that high health expenditures are usually implies improved health care capacities with better monitoring and registrations systems as well as COVID-19 tests. This, consequently, means more recorded cases compared to countries with lower health expenditure.

At the same time, COVID fatality was found to be positively related to asthma prevalence (β=0.00420) and tobacco use (β=0.00005). Yet, certain level of inconsistency was noted in the case of COVID-19 fatality, which was negatively related to elder population (β=-0.00258), poverty (β=-0.00028) and cardiovascular fatality (β=-0.00031). The negative relationship between cardiovascular and COVID-19 fatality can be explained by inaccurate data on cardiovascular fatality due to inefficient national mortality registration systems in most African countries (Rao et al., 2004). Meanwhile, the positive relationship between poverty and COVID-19 fatality can be justified by the fact that poverty usually implies lack of health insurance and less chance of getting hospitalized in case of infection, and less likely to be registered as a Covid-19 death incident.

Moreover, all explanatory variables have noticeably small Variance Inflation Factor (VIF) values, which indicated that no redundancy among the considered explanatory variables. Generally, the results of OLS model are promising as the model recorded R^2^ of 0.4535 in case of COVID-19 incidence and 0.5338 in case of fatality. This means that the model explained about 45% and 53% of the variance in COVID-19 incidence and fatality rates, respectively. However, such a relatively low level of explanatory power of the model can be justified by data accuracy and the nature of OLS itself that doesn’t consider the local variations in COVID-19 incidence and fatality and predictor variables. Accordingly, to increase the explanatory power of the model, Geographically Weighted Regression (GWR) was applied considering spatial relationships between COVID-19 incidence as well as fatality rates and various predictors variables.

The results of GWR were statistically more significant compared to OLS model, where the local produced R^2^ in case of COVID-19 incidence ranged between 0.45 and 0.66 with an overall adjusted R^2^ of 0.58. Meanwhile, local produced R^2^ in case of COVID-19 fatality, was found to be more varied ranging between 0.34 and 0.85 with an overall adjusted R^2^ of 0.55. This is highlighted by the standard variation, which is twofold that of incidence rate (Table 5). Generally, it should be noted that except for BCG vaccine, mostly the GWR model parameters were found to be less varied recording low standard deviation.

**Table 5:** Coefficients, residual and R^2^ produced by (GWR) model

| Parameters | Minimum | Maximum | Mean | Standard Deviation |
| --- | --- | --- | --- | --- |
| COVID-19 incidence |  |  |  |  |
| Intercept | 0.28453 | 1.03252 | 0.58772 | 0.29922 |
| Overcrowding ($\beta$) | 1.98344 | 4.08228 | 3.10461 | 0.49437 |
| Health expenditure ($\beta$) | 0.19844 | 3.33230 | 1.66191 | 1.16584 |
| BCG vaccine ($\beta$) | -128.33056 | -20.09850 | -66.60410 | 42.39930 |
| HIV infections ($\beta$) | 0.00539 | 0.01291 | 0.00986 | 0.00224 |
| Air pollution ($\beta$) | 3.06844 | 4.50561 | 3.78995 | 0.35991 |
| Residual | -0.46329 | 0.47701 | -0.00360 | 0.24063 |
| Local R<sup>2</sup> | 0.44866 | 0.66377 | 0.56400 | 0.06736 |
| Squared residuals | 2.432547 |  |  |  |
| Overall Adjusted R<sup>2</sup> | 0.575972 |  |  |  |
| COVID-19 fatality |  |  |  |  |
| Intercept | -0.00027 | 0.02214 | 0.01078 | 0.00644 |
| Elder population ($\beta$) | -0.00354 | 0.00137 | -0.00144 | 0.00117 |
| Poverty ($\beta$) | -0.00038 | -0.00016 | -0.00025 | 0.00008 |
| Cardiovascular fatality ($\beta$) | -0.00043 | -0.00017 | -0.00027 | 0.00007 |
| Asthma prevalence ($\beta$) | 0.00162 | 0.00625 | 0.15386 | 0.00366 |
| Tobacco use ($\beta$) | 0.00004 | 0.00006 | 0.00005 | 0.00001 |
| Residual | -0.01115 | 0.01649 | 0.00010 | 0.00589 |
| Local R<sup>2</sup> | 0.33673 | 0.85291 | 0.56072 | 0.14392 |
| Squared residuals | 0.00146 |  |  |  |
| Overall Adjusted R<sup>2</sup> | 0.54622 |  |  |  |

Also, it was noted that GWR model in case of COVID-19 incidence revealed higher explanatory power in western and southern parts of Africa compared to other parts of the continent (Figure 4-a). In contrast, higher explanatory power of GWR model in case of COVID-19 fatality was noted in the north-eastern part of Africa and decreases noticeably westward and southward (Figure 4-b).

**Figure 4:**
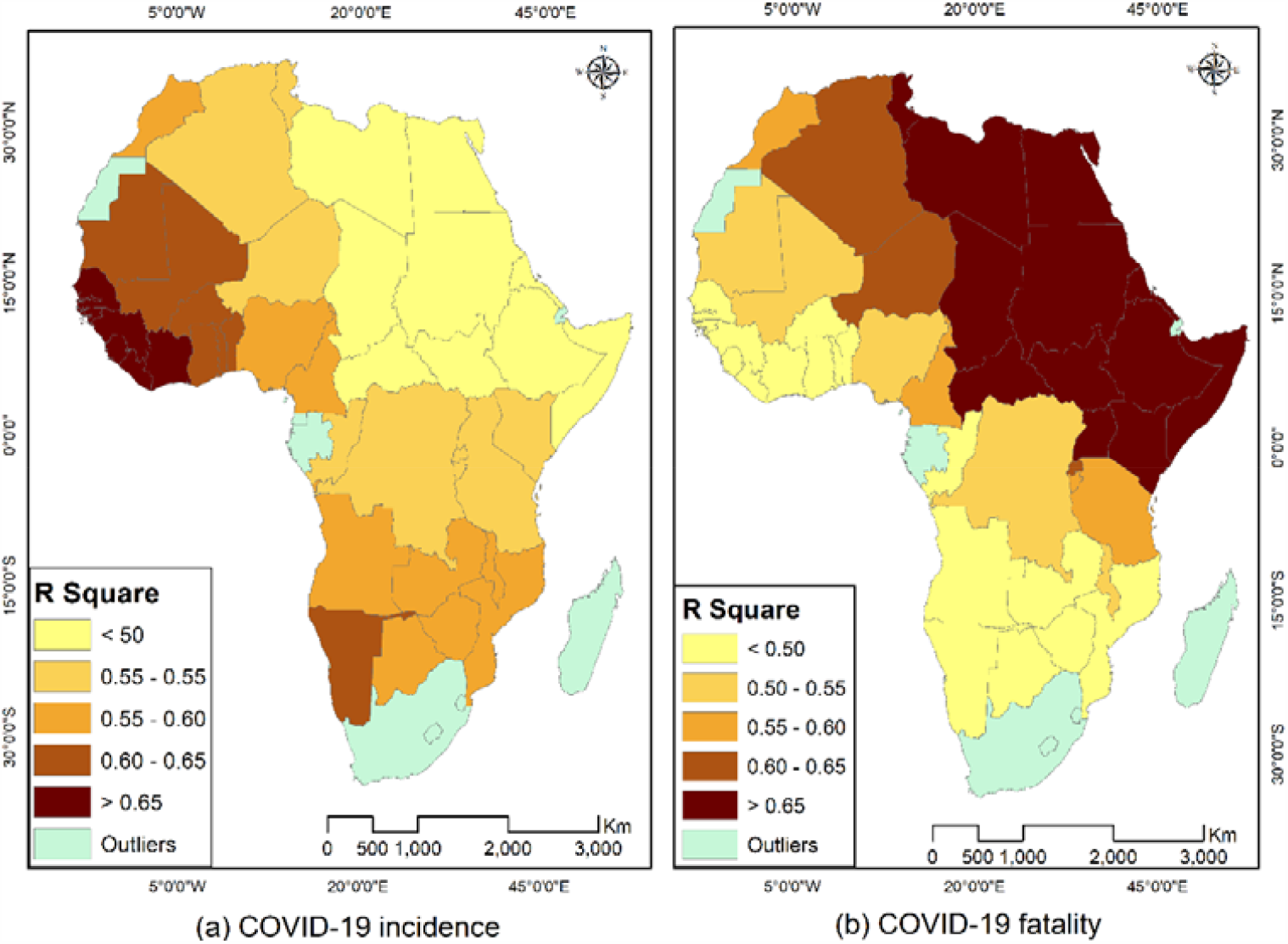
Spatial distribution of local R^2^ of COVID-19 incidence and fatality rates

This means that the relationships between COVID-19 incidence and overcrowding, BCG vaccine, HIV infection, health expenditure and air pollution can be more accurately captured in West Africa and sub-Saharan countries. Meanwhile, the relationships between COVID-19 fatality and elder population, poverty, cardiovascular fatality, asthma prevalence and tobacco use can be more accurately captured in north-eastern African countries.

The validity of GWR model, results was emphasized by the low value of the squared residuals, which was found to be 2.432547 and 0.00146 in the case of COVID-19 incidence and fatality in Africa, respectively. Such a noticeably small value of squared residuals indicates slight difference between observed variable and its estimated value by GWR model and thus model validity. Generally, GWR model revealed varied levels of accuracy in different African countries. This was highlighted by mapping residual values (Figure 5).

**Figure 5:**
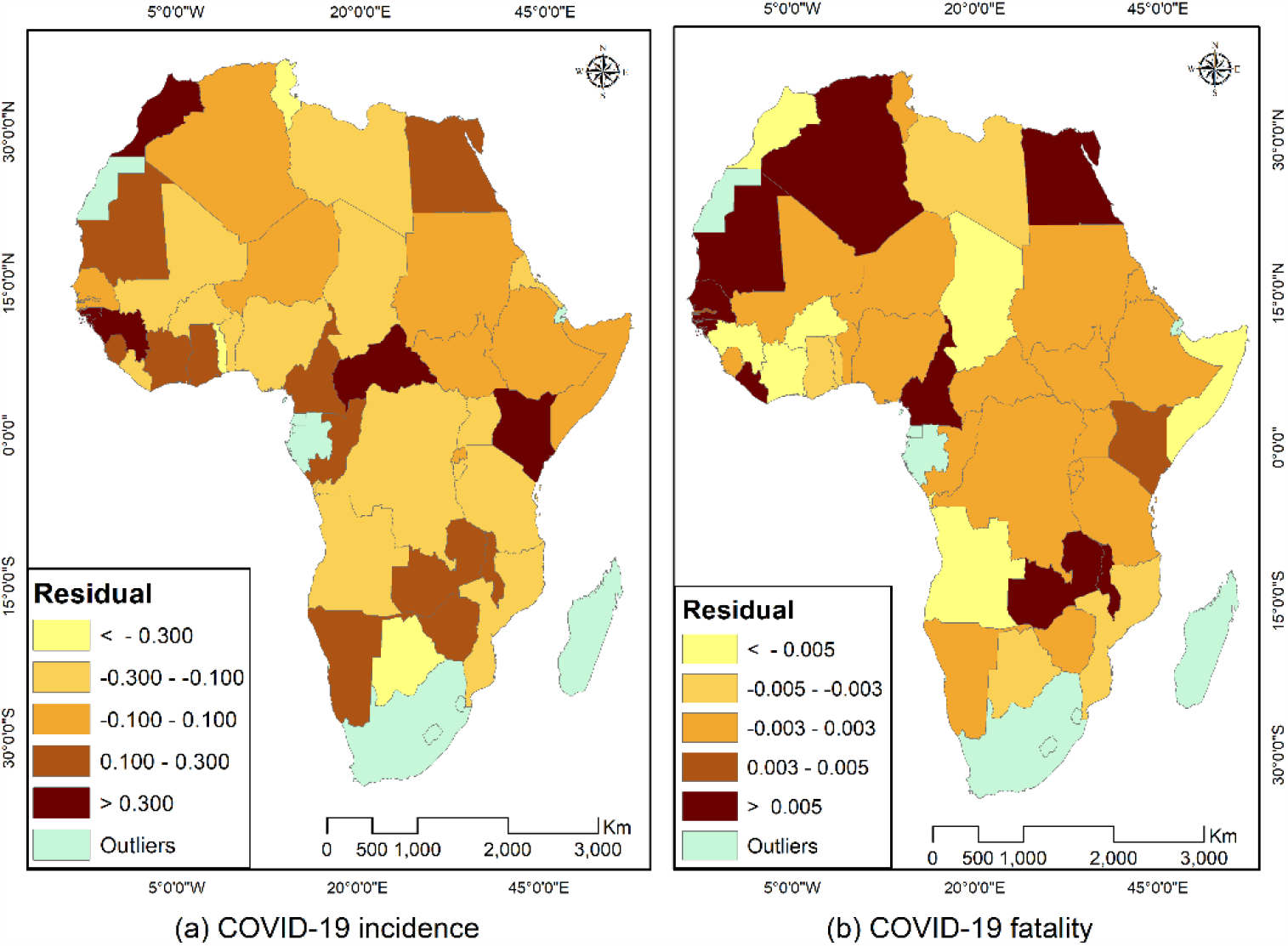
Spatial distribution of residual

Moreover, GWR model predictions were evaluated through plotting COVID-19 incidence and fatality rate in different African countries predicted by the suggested GWR model versus observed rates (Figure 6). In this respect, the suggested GWR model revealed reasonable level of accuracy.

**Figure 6:**
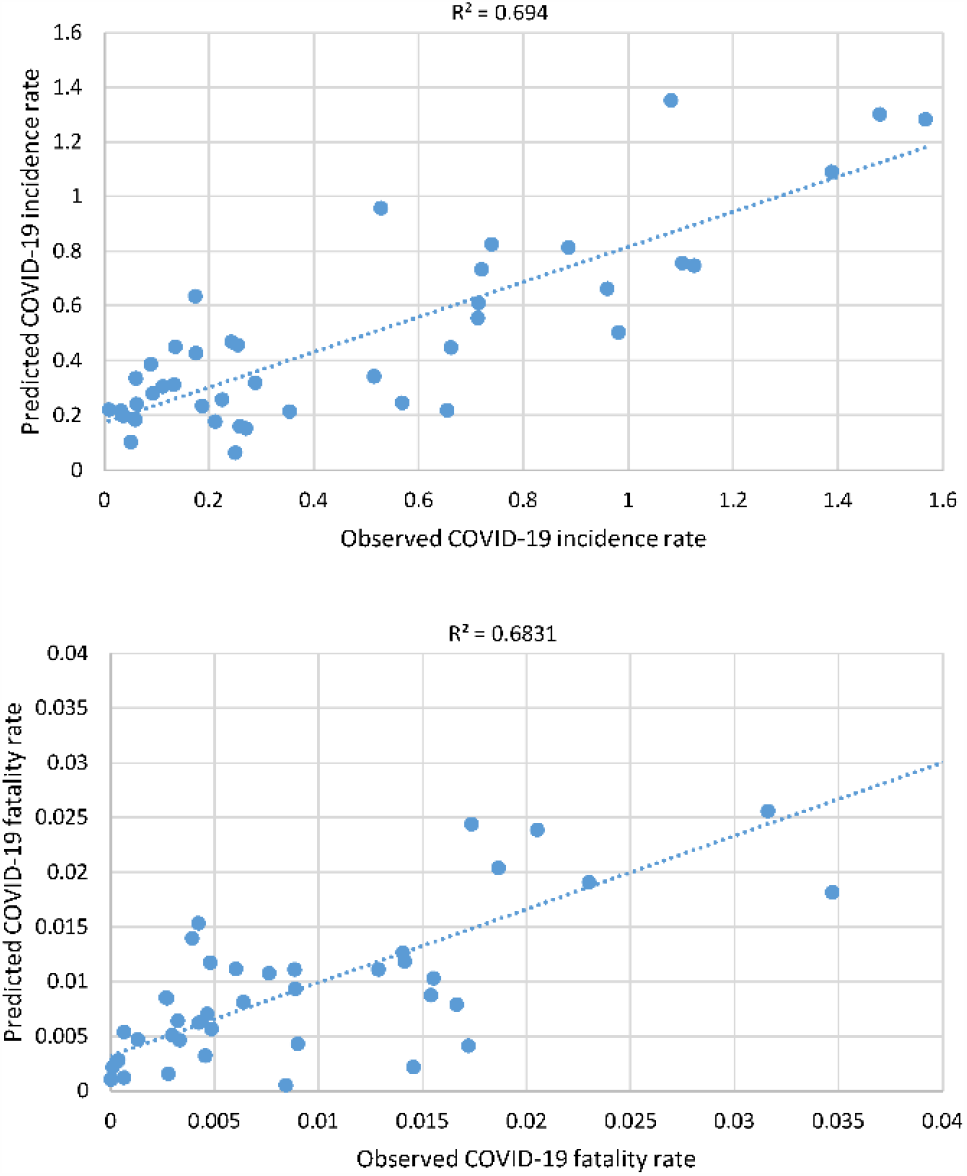
Observed versus predicted COVID-19 incidence and fatality rates in Africa

## Conclusion

The proposed GIS-based model showed high level effectiveness in exploring and modeling the relationships between COVID-19 incidence and fatality rates, on one hand, as dependent variables and a number of relevant predictors as independent variables rate, on the other hand. Modeling such relationships requires using (OLS) and (GWR) in an integrated manner. The relationships between the COVID-19 incidence as well as fatality rates in Africa and various predictor variables have been quantitatively demonstrated through the proposed model.

The proposed model in this study suggested that overcrowding, vaccine against tuberculosis, HIV infection, health expenditure and air pollution are the key determinants of COVID-19 incidence in Africa. Similarly, elder population, poverty, cardiovascular fatality, asthma prevalence and tobacco use were identified through the proposed model to be the key determinants of COVID-19 fatality rate in Africa. Finally, the proposed model can be downscaled to be applied to local and/or community levels. in such a case, the successful application of the GIS-based model suggested in this study depends largely on availability of updated, accurate and detailed data on COVID-19 incidence and fatality as well as various considered predictors.

## Data Availability

Available on request

